# Metabolomics Signatures of serotonin reuptake inhibitor (Escitalopram), serotonin norepinephrine reuptake inhibitor (Duloxetine) and Cognitive Behavior Therapy on Key Neurotransmitter Pathways in Major Depressive Disorder

**DOI:** 10.1101/2024.04.02.24304677

**Authors:** Sudeepa Bhattacharyya, Siamak MahmoudianDehkordi, Matthew J Sniatynski, Marina Belenky, Vasant R. Marur, A. John Rush, W. Edward Craighead, Helen S. Mayberg, Boadie W. Dunlop, Bruce S Kristal, Rima Kaddurah-Daouk, Mood Disorder Precision Medicine Consortium

**Affiliations:** Department of Biological Sciences, Arkansas Biosciences Institute, Arkansas State University, Jonesboro, AR, United States; Department of Psychiatry and Behavioral Sciences, Duke University, Durham, NC, United States; Division of Sleep and Circadian Disorders, Department of Medicine, Brigham and Women’s Hospital, 221 Longwood Ave, LM322B, Boston, MA 02115, USA and Division of Sleep Medicine, Department of Medicine, Harvard Medical School, Boston, MA 02115, USA; Duke-National University of Singapore, Singapore, Singapore; Department of Psychiatry and Behavioral Sciences, Emory University School of Medicine, Atlanta, GA, United States; Department of Neurology and Neurosurgery, Icahn School of Medicine at Mount Sinai, New York, United States; Department of Medicine, Duke University, Durham, NC, United States; Duke Institute of Brain Sciences, Duke University, Durham, NC, United States

## Abstract

Metabolomics provides powerful tools that can inform about heterogeneity in disease and response to treatments. In this study, we employed an electrochemistry-based targeted metabolomics platform to assess the metabolic effects of three randomly-assigned treatments: escitalopram, duloxetine, and Cognitive Behavior Therapy (CBT) in 163 treatment-naïve outpatients with major depressive disorder. Serum samples from baseline and 12 weeks post-treatment were analyzed using targeted liquid chromatography-electrochemistry for metabolites related to tryptophan, tyrosine metabolism and related pathways. Changes in metabolite concentrations related to each treatment arm were identified and compared to define metabolic signatures of exposure. In addition, association between metabolites and depressive symptom severity (assessed with the 17-item Hamilton Rating Scale for Depression [HRSD_17_]) and anxiety symptom severity (assessed with the 14-item Hamilton Rating Scale for Anxiety [HRSA_14_]) were evaluated, both at baseline and after 12 weeks of treatment.

Significant reductions in serum serotonin level and increases in tryptophan-derived indoles that are gut bacterially derived were observed with escitalopram and duloxetine arms but not in CBT arm. These include indole-3-propionic acid (I3PA), indole-3-lactic acid (I3LA) and Indoxyl sulfate (IS), a uremic toxin. Purine-related metabolites were decreased across all arms. Different metabolites correlated with improved symptoms in the different treatment arms revealing potentially different mechanisms between response to antidepressant medications and to CBT.

## INTRODUCTION

Major depressive disorder (MDD) is a serious, disabling, and common condition that is biologically and etiologically heterogeneous[1, 2]. Clinical practice guidelines recommend first step treatment with either an antidepressant medication or an evidence-based psychotherapy, such as cognitive behavior therapy (CBT)[3–5]. Among first-line recommended medications are selective serotonin reuptake inhibitors (SSRIs) and serotonin norepinephrine reuptake inhibitors (SNRIs). Substantial evidence indicates that among outpatients with MDD, medications work as the second step when therapy fails as the first step, and vice versa[6, 7]. Similarly, there is strong evidence that a switch to different medication following a first failed medication trial is likely to be quite effective, demonstrating that mechanistic differences between antidepressants are important to characterize to enable more personalization of treatment [6, 8]. What works well for one patient may not be effective for another, but personalized approaches to MDD treatment are few, resulting in the common trial-and-error approach to treatment selection. Clinicians face a significant challenge in selecting the right treatment for an individual, given the variability in response to antidepressants and psychotherapy. These treatments take weeks to show noticeable benefits[9], which can be discouraging for patients and antidepressants can have side effects ranging from mild to severe[10], impacting a patient’s quality of life and willingness to continue treatment. Furthermore, a significant proportion of patients with MDD do not respond to first-line treatments, necessitating alternative strategies that may include combination therapy, augmentation strategies, or other forms of less traditional treatments[11]. Thus, despite progress in understanding depression and the availability of various treatment options, accurately predicting the most effective treatment for individual patients remains a significant challenge. Additionally, the specific molecular mechanisms that underlie the effectiveness of CBT, how these mechanisms differ from those involved in medication treatments, and why CBT may be more effective for certain patients are not well understood. Collectively, these insights indicate that treatment strategies for MDD, including both CBT and medication, may achieve remission through biologically distinct pathways.[12–14].

We previously have shown that metabolomic profiling is a promising approach that can provide informative insights into the molecular mechanisms of h SSRIs and how they differentially impact the metabolome in patients with MDD[15, 16]. In addition, we have provided insights into rapid metabolomic effects of ketamine[17, 18]. Prior findings suggested that in patients with MDD, the tryptophan pathway and its branches leading to serotonin, melatonin, 5-hydroxyindoleacetate, kynurenine, and indole derivatives appear to be significantly altered compared to healthy controls [15, 19]. The purine pathway, whose regulation is linked to tryptophan metabolism, has also been connected to MDD and other psychiatric illnesses[20]. Branched chain amino acids (BCAAs) that are linked to short chain acylcarnitine biosynthesis and mitochondrial energetics have been implicated in depression in several recent studies[21, 22]. Lower blood levels of BCAAs were associated with depression and increase in their level through dietary intake associated with improved symptoms[23]. However, no randomized control trial studies to our knowledge have directly compared the effects of treatment with psychotherapy versus one or more antidepressant medications on the metabolic pathways of neurotransmitters commonly implicated in MDD, such as serotonin, norepinephrine, or dopamine among MDD patients.

In this report, we used a highly sensitive quantitative electrochemistry-based platform to interrogate effect of 12 weeks’ of treatment on key neurotransmitter pathways, including tryptophan, tyrosine and related pathways, among MDD patients randomly assigned to receive escitalopram (an SSRI), duloxetine (an SNRI), and CBT in the Predictors of Remission in Depression to Individual and Combined Treatments (PReDICT) study[24]. This secondary data analysis examined the following aims:

1. Identify baseline metabolites associated with baseline depressive symptoms (assessed with the 17-item Hamilton Rating Scale for Depression [HRSD_17_][25]) severity and baseline anxiety (assessed with the 14-item Hamilton Rating Scale for Anxiety [HRSA_14_][26]) severity.
2. Define metabolic signatures for treatment with escitalopram, duloxetine, and CBT; Compare and contrast these signatures.
3. Define metabolic changes associated with changes in depressive symptoms (HRSD_17_) and changes in anxiety (HRSA_14_) over 12 weeks of treatment within each treatment arm.

## 2. MATERIALS AND METHODS

### 2.1 Study Design and Participants

The design and clinical outcomes of PReDICT have been detailed previously[7, 24, 27]. The PReDICT study was conducted through the Mood and Anxiety Disorders Program of Emory University from 2007-2013 (Clinical trial NCT00360399 (08/04/2006) “Predictors of Antidepressant Treatment Response: The Emory CIDAR” https://clinicaltrials.gov/ct2/show/NCT00360399). The study was approved by the Emory Institutional Review Board. All patients provided written informed consent to participate. The study was conducted in accordance with the 1975 Declaration of Helsinki and its amendments. Briefly, PReDICT aimed to identify predictors and moderators of response to 12 weeks of randomly assigned treatment with duloxetine (30-60 mg/day), escitalopram (10-20 mg/day) or CBT (16 one-hour individual sessions). Eligible participants were adults aged 18-65 with non-psychotic MDD who had never previously been treated for depression. The diagnosis of MDD was made via interview by assessors trained in administering the Structured Clinical Interview for DSM-IV (SCID)[28]. The SCID was also used to diagnose any comorbid psychiatric disorders, including the exclusionary diagnoses of bipolar disorder, psychotic disorder, obsessive compulsive disorder, anorexia nervosa, and current substance abuse or dependence. Additional exclusionary criteria included a neurocognitive disorder, pregnancy, lactation, any uncontrolled general medical condition, or a positive urine drug screen for illicit drugs. Severity of depression at the randomization visit was assessed with the HRSD_17_. Eligibility required an HRSD_17_ score ≥18 at the screening visit and ≥15 at the randomization visit, indicative of moderate-to-severe depression.

### 2.2 Metabolomic Profiling and Ratios

On the day of randomization (without consideration of diet, time of day or fasting status), 10 cc of whole blood was collected in a red-top silicone-coated tube from an antecubital vein between 8 am and 4 pm. After the blood had clotted for 20 min, the tubes were centrifuged at 1400 g at 4LC for 10 mins. Serum was then pipetted into 1 mL vials and frozen at −80 C until thawed for analyses.

#### 2.2.1 High Performance Liquid Chromatograph-Electrochemical detection (HPLC-ECD)

Metabolite extraction from human serum and analyses by HPLC-ECD were conducted as described in detail previously,14-20 with minor changes (e.g., specific columns) as noted below. Briefly, the proteins from 125 μL of serum were precipitated by adding 0.5 ml of cold (−20°C) acetonitrile/0.4% glacial acetic acid. After centrifugation for 15 minutes at 4°C, at 15000 x g, the supernatant was transferred into microcentrifuge tubes and dried under vacuum. The dried extract was reconstituted in 100 μL of mobile phase A prior to injection; 50 μL of each sample was injected.

Reversed phase LC separation was performed using buffers containing high concentration of salts and pentane sulfonic acid (PSA). Salts were beneficial to maintain conductivity of the EC electrodes, and PSA was used as an ion-pairing reagent to help the retention and separation of hydrophilic metabolites. In this study, we used our standard serum metabolites profiling LC separation method without any modifications to see which 2-oxoacids could be quantified along with other redox metabolites we routinely analyze. Two connected reversed-phase C18 Luna 250 × 4.6 × 5 μm columns (Phenomenex, Torrance, CA) at 30°C were used to improve separation. Mobil phase A was comprised of 60mM aqueous solution of sodium 1-pentasulfonate (PSA), 0.1% methanol, and 1mg/L citric acid. Mobile phase B consisted of 80:10:10 MeOH/ACN/IPA with 40 mM lithium acetate, 2.0% acetic acid, and 10 mg/L citric acid. The separation was completed over 112 minutes.

Detection was done using 16-channel coulometric array detector (i.e., CoulArrayTM, combining four of Hi-E four sensors assembly cells, (ThermoFisher Scientific, Chelmsford, MA) operated with potentials incremented in 60 mV steps (0−900 mV). In our system, Pd electrode is used as a reference electrode. The HPLC-ECD system was controlled by CoulArray software (CoulArray software for Windows, version 3.10). Data analysis was performed using CoulArray Data Station software for Windows.

#### 2.2.2 Biocrates P180 Platform

Branched-chain amino acids valine and isoleucine were quantified with a targeted metabolomics approach using the AbsoluteIDQ® p180 Kit (BIOCRATES Life Science AG, Innsbruck, Austria), with an ultra-performance liquid chromatography (UPLC)/MS/MS system [Acquity UPLC (Waters), TQ-S triple quadrupole MS/MS (Waters)]. This platform provides measurements of up to 186 endogenous metabolites in quantitative mode (amino acids and biogenic amines) and semi-quantitative mode (acylcarnitines, sphingomyelins, phosphatidylcholines and lysophosphatidylcholines across multiple classes). The AbsoluteIDQ® p180 kit has been fully validated according to European Medicine Agency Guidelines on bioanalytical method validation. The p180 kit includes all quality control (QC) samples, and calibration and internal standards; therefore, quantifications can be directly compared across studies. The quality-control and metabolomic profiling were previously published[16, 29, 30]. In short, metabolites with 1) > 40% of measurements below the lower limit of detection (LOD) and 2) > 30% of SPQC coefficient of variation were excluded from the analysis. To adjust for the batch effects, a correction factor for each metabolite in a specific plate was obtained by dividing the metabolite’s QC global average by QC average within the plate. Missing values were imputed using each metabolite’s LOD/2.

#### 2.2.3 Metabolic Ratios

To determine which enzymatic processes may be affected by the disease and/or treatment, we expanded metabolic profile with certain ratios of related metabolites within tryptophan, tyrosine, purine and BCAA pathways. These ratios included:

1. Purine Pathway: Uric:Xanthine, Hypoxanthine: Xanthine, Hypoxanthine:Uric
2. Tryptophan Pathway: Tryptophan:Tyrosine, Indole-3-acetic acid:Tryptophan, Indole-3-lactic acid:Tryptophan, Indole-3-propionic acid:Tryptophan, Indoxyl sulfate: Tryptophan, Indoxyl sulfate: Indole-3-propionic acid, 5HT: Tryptophan, Tryptophan: Kynurenine
3. BCAA Pathway: Three ratios were tested: 1) 4-methyl-2-oxopentanoate (b036):Isoleucine, 2) b036:valine and 3) the ratio of b036:3-methyl-2-oxopentanoate (b032).

### 2.3 Depression and Anxiety Symptoms

At baseline and after 12 weeks of treatment, severity of depression and anxiety was assessed by blinded raters using the HRSD_17_ and HRSA_14_, respectively.

### 2.4 Statistical Analysis

Differences in demographic variables and depression scores across the response groups were evaluated using ANOVA and the Pearson Chi-squared test (for categorical variables). Patients with body mass index (BMI) ≥ 30 were labeled as obese vs. BMI<30 as “Non-obese”. All analyses were performed in a metabolite-wise manner in 1) all samples, 2) males, 3) females, 4) obese, 5) non-obese, 6) Hispanic, and 7) non-Hispanic.

To assess the significance of a metabolite’s log_2_ fold-change within each subpopulation, bootstrapping with 1000 repeats were conducted using the “bootstraps” function from the “rsample” package. To assess the association of baseline metabolite levels and the continuous variables HRSD_17_ and HRSA_14_, partial correlation analyses were conducted using partial Spearman rank correlation, adjusted for age, sex, and body mass index. Significance of partial correlation were computed using bootstrapping with 1000 repeats. A p-value <0.05 was considered significant. Given the exploratory nature of this initial investigation, no correction for multiple comparisons was made.

## 3. RESULTS

### 3.1 Participant Characteristics (Demographic and Clinical)

**Table 1** summarizes the demographic and clinical features of the 163 participants in the PReDICT Study. Of these, 41% of participants were male, and mean (standard deviation) baseline age, HRSD_17_, and HRSA_14_ were 39(12) years, 20(3.8) points, and 16 (5.2) points, respectively. Of 163 participants, 131 individuals had both baseline and week 12 electrochemistry-based profile. and 107 individuals had valine and isoleucine measurement at both visits as these two compounds were profiled on the p180 platform. Baseline total HRSD_17_ scores were highly correlated with HRSA_14_ scores (Spearman rank correlation rho= 0.61, p = 2.2E-16).

**Table 1:** Participant Demographic and Clinical Characteristics.

| Variable | Baseline |  | Week 12 |  | Test |
| --- | --- | --- | --- | --- | --- |
|  | N | Mean(SD) or % | N | Mean(SD) or % |  |
| **Age** | 163 | 39(12) | 131 | 39(12) |  |
| **Sex** | 163 |  | 131 |  |  |
| **Female** | 97 | 60% | 78 | 60% |  |
| **Male** | 66 | 40% | 53 | 40% |  |
| **Baseline BMI** | 163 | 29(6.4) | 131 | 29(6.6) |  |
| **Hispanic** | 163 |  | 131 |  |  |
| **Yes** | 59 | 36% | 42 | 32% |  |
| **No** | 104 | 64% | 89 | 68% |  |
| **White, Black Non-Hispanic** | 163 |  | 131 |  |  |
| **Yes** | 93 | 57% | 80 | 61% |  |
| **No** | 70 | 43% | 51 | 39% |  |
| **Baseline Obese** | 163 |  | 131 |  |  |
| **Non-Obese (BMI<30)** | 99 | 61% | 81 | 62% |  |
| **Obese (BMI>30)** | 64 | 39% | 50 | 38% |  |
| **Treatment** | 163 |  | 131 |  |  |
| **CBT** | 49 | 30% | 37 | 28% |  |
| **Duloxetine** | 52 | 32% | 44 | 34% |  |
| **Escitalopram** | 62 | 38% | 50 | 38% |  |
| **HRSD<sub>17</sub>** | 163 | 20(3.8) | 129 | 7.2(6.1) | F=457.318*** |
| **HRSA<sub>14</sub>** | 163 | 16(5.2) | 128 | 6.6(5.8) | F=214.93*** |

*Abbreviations:* HRSA<sub>14</sub>: 14-item Hamilton Anxiety Rating Scale. HRSD<sub>17</sub>: 17-item Hamilton Depression Rating Scale.

### 3.2 Baseline Profiles and Disease Severity

We explored metabolite profiles at baseline that correlated with depression and anxiety symptom severity (i.e., HRSD_17_, and HRSA_14_) using Spearman partial correlation analysis adjusted for age, sex, and BMI. Estimated partial correlations (rho) and *p*-values were obtained from bootstrapping and are presented in **Table 2** for the overall samples and in **Supplementary Fig 1** for stratified analyses. We identified 14 metabolites and ratios at baseline that were significantly correlated with depression severity and/or anxiety severity. As expected, due to a high correlation between these two outcomes (rho=0.61), the direction of correlations was similar.

**Table 2:** Significant Spearman Partial Correlation of Baseline Metabolites/Ratios and Disease Severity. Spearman Partial correlations were adjusted for age, sex, and baseline BMI. Partial correlations, p-values and 95% confidence intervals were calculated using bootsrtaping with 1000 repetitions. *: p<0.05; **: p<0.01; ***: p<0.001

| Abbreviation | Full name | Pathway | Partial Correlation (95% Bootstrap CI)<br>p-value |  |
| --- | --- | --- | --- | --- |
|  |  |  | HRSD<sub>17</sub> | HRSA<sub>14</sub> |
| IS | Indoxyl sulfate | Tryptophan metabolism | 0.10<br>(-0.05,0.25)<br>p>0.05 | 0.22<br>(0.07,0.36)<br>** |
| KYN | Kynurenine | Tryptophan metabolism | 0.08<br>(-0.08,0.23)<br>p>0.05 | 0.23<br>(0.08,0.37)<br>** |
| TRP | Tryptophan | Tryptophan metabolism | 0.20<br>(0.04,0.34)<br>* | 0.17<br>(0.02,0.31)<br>* |
| HVA | Homovanillic acid | Phenylalanine/Tyrosine metabolism | -0.08<br>(-0.23,0.08)<br>p>0.05 | -0.16<br>(-0.30,-0.00)<br>* |
| TYR | Tyrosine | Phenylalanine/Tyrosine metabolism | 0.20<br>(0.04,0.35)<br>** | 0.21<br>(0.07,0.35)<br>** |
| Cystine | Cystine | Cysteine/Methionine metabolism | 0.19<br>(0.03,0.33)<br>* | 0.24<br>(0.08,0.38)<br>*** |
| MET | Methionine | Cysteine/Methionine metabolism | 0.16<br>(0.01,0.31)<br>* | 0.13<br>(-0.02,0.28)<br>p>0.05 |
| SAH | S-(5'-Adenosyl) homocysteine | Cysteine/Methionine metabolism | 0.09<br>(-0.12,0.29)<br>p>0.05 | 0.22<br>(0.04,0.42)<br>* |
| Pyruvate+ | Pyruvate and other metabolites, like oxaloacetic acid are measured together | Glycolysis | 0.14<br>(-0.02,0.28)<br>p>0.05 | 0.27<br>(0.13,0.41)<br>*** |
| b032 | 3-methyl-2-oxopentanoate | BCAA biosynthesis/metabolism | 0.19<br>(0.03,0.34)<br>** | 0.18<br>(0.03,0.32)<br>* |
| b036 | 4-methyl-2-oxopentanoate | BCAA biosynthesis/metabolism | 0.20<br>(0.04,0.36)<br>* | 0.17<br>(0.02,0.32)<br>* |
| 3HHA | 3-hydroxyhippuric acid | Other | -0.22<br>(-0.37,-0.08)<br>** | -0.12<br>(-0.28,0.03)<br>p>0.05 |
| I3PA:TRP |  | Tryptophan Ratio | -0.15<br>(-0.29,-0.00) | -0.04<br>(-0.20,0.12) |
|  |  |  | * | p>0.05 |
| IS:TRP |  | Tryptophan Ratio | 0.04<br>(-0.13,0.19)<br>p>0.05 | 0.16<br>(0.00,0.31)<br>* |

In *the tryptophan pathway*, tryptophan showed a positive correlation with both depression and anxiety symptom severity, while indoxyl sulfate and kynurenine were only significantly positively correlated with anxiety symptom.

In *the phenylalanine/tyrosine pathway*, tyrosine showed positive correlations with both depression and anxiety symptoms similar to tryptophan. Higher levels of homovanillic acid were associated with less severe anxiety symptoms.

In *the cysteine/methionine pathway*, cystine, an oxidized dimeric form of the amino acid cysteine, was significantly positively associated with greater severity of both depression and anxiety symptoms; methionine was positively associated with depression symptoms and higher levels of S-adenosyl-homocysteine, which is a precursor of all homocysteine, was associated with greater anxiety symptoms.

In *the glycolysis/energy metabolism pathway*, higher levels of pyruvate showed a significant positive correlation with higher anxiety symptoms.

3-methyl-2-oxopentanoate and 4-methyl-2-oxopentanoate, two metabolites of the BCAAs valine and isoleucine, were also positively correlated with more severe depression and anxiety scores.

The ratio of I3PA:TRP showed a negative correlation with depression severity and IS:TRP showed a positive significant correlation with total anxiety score, suggesting decreased production of I3PA and increased production of indoxyl sulfate from tryptophan in patients with more severe symptoms.

### 3.3 Metabolite Signatures of 12 Weeks of treatment with antidepressant medication or CBT

All three treatments triggered significant changes in metabolites implicated in neurotransmitter related pathways. However, when compared to CBT, the changes observed in the medication arms were more similar **(Figure 1).** In addition, we noted similar signatures across different strata (men, women, Hispanic, non-Hispanic, obese, non-obese) of the study (**Supplementary Fig 2**).

**Figure 1:**
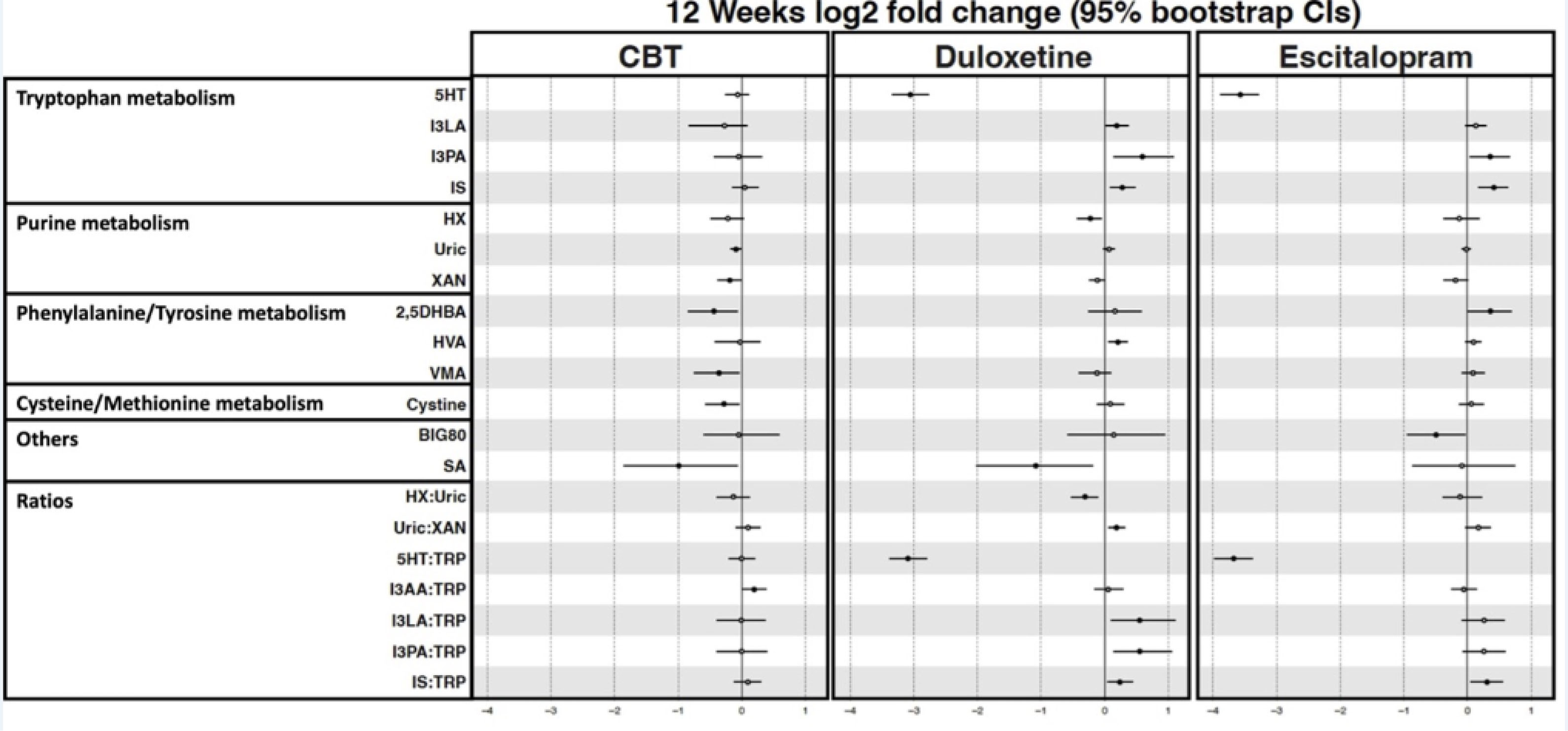
Signature of Exposure. Forest plot shows 12 weeks log_2_ fold change and 95% bootstrap confidence intervals within each treatment group. P-values were computed using bootstrapping with 1000 repetitions. A negative value represents decrease in metabolite levels and a positive value shows an increase.

*<u>In the tryptophan pathway</u>*, the largest impact was observed in serotonin levels which decreased significantly in both the escitalopram and duloxetine arms, while no change was observed in the CBT arm. The ratio of serotonin to tryptophan was also significantly lower in the medication arms but remained unchanged in CBT. The indole containing metabolites of tryptophan, primarily generated by the gut bacteria such as indoxyl sulfate, indole 3-lactic acid and indole 3-propionic acid were all increased by medications but not CBT. The ratios of each of the indoles to tryptophan were also higher in the medication arms but showed no change in CBT.

*<u>In the purine pathway,</u>* hypoxanthine, xanthine and uric acid decreased or trended to decrease in all three arms. Specifically, uric acid and xanthine in the CBT arm, hypoxanthine in the duloxetine arm, and xanthine in the escitalopram arm showed significant decreases with treatment.

*<u>In the phenylalanine/tyrosine pathway</u>*, homovanillic acid showed small increases in the medication arms, which was statistically significant with duloxetine. Vinylmandelic acid, a metabolic by-product of norepinephrine and epinephrine, showed significant decrease in the CBT arm but not in the medication arms.

Among other molecules, salicylic acid (SA) and its active metabolite 2,5-dihydroxybenzoic acid, also known as gentisic acid, showed significant decreases in the CBT arm. SA was also decreased with exposure in the duloxetine arm but not in the escitalopram arm.

### 3.3 Metabolic system changes in relation to depressive and anxious symptom change for each treatment

We compared the metabolite changes that correlated with improved symptoms of depression and anxiety distress upon treatment with medications and CBT treatments. **Table 3** lists the metabolites that showed changes in their serum levels associated with changes in symptom severity.

**Table 3:** Significant Associations Between %Change in Disease Severity Score and Log2 Fold Change Metabolites/Ratios. Linear regression models were adjusted for age, sex, and baseline BMI. *p*-values and 95% confidence intervals were calculated using bootstrapping with 1000 repetitions. Positive beta values indicate that decreases in metabolites or ratios changes were associated with improvement in symptoms; negative beta values indicate that increases in metabolites or ratios were associated with improvement in symptoms. Shaded boxes reflect significant associations.*: p<0.05; **: p<0.01; ***: p<0.001

| Abbreviation | Full name | Pathway | Beta (95% Bootstrap CI);p-value |  |  |  |  |  |
| --- | --- | --- | --- | --- | --- | --- | --- | --- |
|  |  |  | ΔMetabolite~ΔHRSD<sub>17</sub> |  |  | ΔMetabolite~ΔHRSA<sub>14</sub> |  |  |
|  |  |  | CBT | Duloxetine | Escitalopram | CBT | Duloxetine | Escitalopram |
| IS | indoxyl sulfate | Tryptophan metabolism | 0.01<br>(-0.54,0.51)<br>*p*>0.05 | -0.92<br>(-1.83,-0.24)<br>* | 0.29<br>(-0.54,1.26)<br>*p*>0.05 | -0.08<br>(-0.55,0.40)<br>*p*>0.05 | -0.44<br>(-1.18,0.16)<br>*p*>0.05 | 0.14<br>(-0.54,0.96)<br>*p*>0.05 |
| KYN+ | kynurenine | Tryptophan metabolism | 0.09<br>(-0.19,0.32)<br>*p*>0.05 | -0.48<br>(-0.82,-0.12)<br>* | 0.12<br>(-0.28,0.52)<br>*p*>0.05 | 0.06<br>(-0.23,0.25)<br>*p*>0.05 | -0.38<br>(-0.68,-0.10)<br>* | 0.07<br>(-0.25,0.38)<br>*p*>0.05 |
| HX | hypoxanthine | Purine metabolism | 0.66<br>(0.05,1.26)<br>* | 0.29<br>(-0.47,0.97)<br>*p*>0.05 | -0.06<br>(-0.73,0.68)<br>*p*>0.05 | 0.49<br>(-0.16,1.17)<br>*p*>0.05 | -0.07<br>(-0.98,0.68)<br>*p*>0.05 | -0.09<br>(-0.78,0.44)<br>*p*>0.05 |
| XAN | xanthine | Purine metabolism | 0.71<br>(0.30,1.04)<br>*** | 0.48<br>(0.05,0.98)<br>*p*>0.05 | -0.28<br>(-0.96,0.45)<br>*p*>0.05 | 0.60<br>(0.20,1.10)<br>* | 0.33<br>(-0.02,0.74)<br>*p*>0.05 | -0.24<br>(-0.87,0.39)<br>*p*>0.05 |
| Ile | Isoleucine | BCAA biosynthesis/metabolism | -0.68<br>(-1.29,0.19)<br>* | -0.32<br>(-0.84,0.33)<br>*p*>0.05 | 0.34<br>(-0.60,1.10)<br>*p*>0.05 | -0.49<br>(-1.13,0.50)<br>*p*>0.05 | -0.36<br>(-0.84,0.13)<br>*p*>0.05 | 0.28<br>(-0.46,1.01)<br>*p*>0.05 |
| Val | Valine | BCAA biosynthesis/metabolism | -0.54<br>(-0.99,-0.10)<br>* | -0.45<br>(-0.85,-0.01)<br>* | 0.29<br>(-0.42,0.88)<br>*p*>0.05 | -0.37<br>(-0.87,0.13)<br>*p*>0.05 | -0.41<br>(-0.78,-0.06)<br>* | 0.26<br>(-0.26,0.76)<br>*p*>0.05 |
| b032 | 3-methyl-2-oxopentanoate | BCAA biosynthesis/metabolism | -0.19<br>(-0.71,0.28)<br>*p*>0.05 | 0.60<br>(0.07,1.19)<br>* | -0.08<br>(-0.55,0.35)<br>*p*>0.05 | -0.05<br>(-0.44,0.39)<br>*p*>0.05 | 0.30<br>(-0.17,0.79)<br>*p*>0.05 | 0.01<br>(-0.38,0.39)<br>*p*>0.05 |
| b036 | 4-methyl-2-oxopentanoate | BCAA biosynthesis/metabolism | -0.10<br>(-0.60,0.27)<br>*p*>0.05 | 0.56<br>(0.06,1.05)<br>* | -0.07<br>(-0.50,0.38)<br>*p*>0.05 | 0.01<br>(-0.31,0.32)<br>*p*>0.05 | 0.15<br>(-0.30,0.59)<br>*p*>0.05 | 0.00<br>(-0.40,0.42)<br>*p*>0.05 |
| 3HHA | 3-hydroxyhippuric acid | Other | 0.22<br>(-3.00,3.44)<br>*p*>0.05 | -2.86<br>(-5.59,-0.21)<br>* | -1.72<br>(-4.29,0.66)<br>*p*>0.05 | -0.76<br>(-4.01,2.45)<br>*p*>0.05 | -2.95<br>(-5.72,-0.45)<br>* | -0.56<br>(-3.04,1.48)<br>*p*>0.05 |
| Ratios |  |  |  |  |  |  |  |  |
| Abbreviation | Full name | Pathway | CBT | Duloxetine | Escitalopram | CBT | Duloxetine | Escitalopram |
| Uric:XAN |  | Purines Ratio | -0.66<br>(-1.05,-0.20)<br>** | -0.58<br>(-1.02,-0.10)<br>* | 0.24<br>(-0.51,1.00)<br>*p*>0.05 | -0.51<br>(-1.12,-0.04)<br>*p*>0.05 | -0.42<br>(-0.83,0.03)<br>*p*>0.05 | 0.22<br>(-0.42,0.88)<br>*p*>0.05 |
| IS:TRP |  | Tryptophans Ratio | 0.08<br>(-0.59,0.60)<br>*p*>0.05 | -0.96<br>(-1.93,-0.15)<br>* | 0.14<br>(-0.67,1.07)<br>*p*>0.05 | 0.05<br>(-0.46,0.56)<br>*p*>0.05 | -0.40<br>(-1.18,0.21)<br>*p*>0.05 | 0.08<br>(-0.67,0.91)<br>*p*>0.05 |
| TRP:KYN |  | Tryptophans Ratio | -0.22<br>(-0.46,0.01)<br>*p*>0.05 | 0.53<br>(0.17,0.95)<br>* | 0.00<br>(-0.43,0.39)<br>*p*>0.05 | -0.21<br>(-0.45,0.07)<br>*p*>0.05 | 0.34<br>(0.02,0.66)<br>* | 0.01<br>(-0.33,0.37)<br>*p*>0.05 |
| b036:Ile |  | BCAAs Ratio | 0.23<br>(-0.59,0.86)<br>*p*>0.05 | 0.79<br>(0.17,1.46)<br>* | -0.53<br>(-1.21,0.27)<br>*p*>0.05 | 0.28<br>(-0.49,0.83)<br>*p*>0.05 | 0.58<br>(0.07,1.09)<br>* | -0.32<br>(-0.92,0.31)<br>*p*>0.05 |
| b036:Val |  | BCAAs Ratio | 0.29<br>(-0.50,0.84)<br>*p*>0.05 | 0.88<br>(0.28,1.53)<br>** | -0.34<br>(-0.99,0.40)<br>*p*>0.05 | 0.26<br>(-0.44,0.72)<br>*p*>0.05 | 0.48<br>(0.00,1.06)<br>*p*>0.05 | -0.24<br>(-0.77,0.37)<br>*p*>0.05 |

Among the metabolites of *<u>tryptophan pathway</u>*, indoxyl sulfate and kynurenine increased with improvements in depression and anxiety symptoms in the duloxetine arm. The ratios of IS:TRP and KYN:TRP were significantly negatively correlated with depression symptom severity in the duloxetine arm.

*<u>In the purine pathway</u>*, Hypoxanthine and xanthine were decreased in the CBT arm and were significantly correlated with improvements in depression and anxiety symptom severity. Xanthine also showed significant positive correlation with depression symptom severity in the duloxetine arm but not in the escitalopram arm. The ratio of Uric acid:Xanthine was increased with improvements in depression symptom severity in both CBT and duloxetine arms.

*<u>The branched-chain amino acid (BCAA),</u>* Increase in valine was significantly associated with improvements in depressive symptom severity both in CBT and duloxetine arms and also correlated with decrease in anxiety symptom severity in the duloxetine arm and showed a similar increasing trends (non-significant at p<0.05) in the CBT arm. In contrast, decrease in two metabolites of the BCAA biosynthesis pathway, 3-methyl-2-oxopentanoate and 4-methyl-2-oxopentanoate were associated with improvements in depressive symptoms in the duloxetine arm. The ratios of 4-methyl-2-oxopentanoate/valine and 4-methyl-2-oxopentanoate/Isoleucine were positively correlated with depressive symptom severity in the duloxetine arm.

## 4. DISCUSSION

In this study, we applied a targeted liquid-chromatography coupled to electrochemical coulometric array (LCECA) metabolomics platform to compare metabolite profiles in serum samples from MDD patients in the three arms of the PreDICT study. The targeted metabolites were amino acids and redox metabolites primarily related to neurotransmitter pathways.

Among the highlights of our findings in the MDD patients was the replication of our metabolomic analyses from other depression samples, that antidepressant medications significantly lowered serum serotonin levels; further, and the ratio of serotonin to its parent metabolite, tryptophan, after 12 weeks of treatment. Our results, for the first time showed that this decrease in serotonin levels was not observed in the CBT arm. We and others[15, 16, 31–33] have shown that plasma 5-HT levels decreased after SSRI treatment, and that higher baseline 5HT levels were associated with better treatment response. The exact mechanism associated with this robust decrease in circulating serotonin with the administration of SSRIs are not known even though several mechanisms including 5-HT1A autoreceptor hypofunctioning[32, 34] and potential SSRI-induced blockade of platelet uptake of 5-HT from cells in the gut[32] have been suggested.

The other major differentiating metabolomic effect of the treatments was the significant shift towards the indole acids branch of the tryptophan metabolic pathway in the medication arms but not in CBT. Indole 3-lactic acid, indole 3-propionic acid and indoxyl sulfate, and their ratios to tryptophan, all increased significantly with 12 weeks of drug exposure, but were not observed in the CBT arm. This shift in the tryptophan metabolic pathway towards increased production of the gut-microbiota-synthesized indoles with exposure to antidepressants has been reported previously by our group in multiple studies[15, 35].

These observations highlight the differences in the mode of action of the drugs and the psychotherapy even though both are efficacious in the treatment and remission of MDD. Indoxyl sulfate is a known uremic toxin that has been shown, by our group, to be associated with increased severity of depression and anxiety symptoms in depression patients[35].

Given these associations, the finding that treatment with either escitalopram or duloxetine, significantly increased the serum levels of indoxyl sulfate was surprising. This increase in uremic toxin can be interpreted as potential side-effect of the drugs via modulation of gut microbiome. More investigations are needed to confirm this increase in indoxyl sulfate as a possible consequence of these antidepressant medications.

Across all three treatments there were statistically significant, or trend-level decreases of purine pathway metabolites, including hypoxanthine, xanthine, and uric acid and they were associated with improved symptoms in CBT and duloxetine arms. These results may indicate that the purine pathway metabolites are associated with or are reflective of treatment response in these patients. This corroborates our previous findings on a different MDD cohort exposed to either citalopram/escitalopram[15] or sertraline (other SSRIs) or placebo[36], wherein we had observed significant decreases or decreasing trends in xanthine and hypoxanthine levels in both drugs or placebo groups, with exposure. It is possible that decreases in these purine metabolites indicate reduction in oxidative stress by direct or indirect inhibition of the xanthine oxidase enzyme that generates vascular oxidative stress through reactive oxygen species production by catalyzing the hypoxanthine to xanthine to urate synthesis pathway.

Branched chain amino acids like valine, leucine and isoleucine have been suggested to play crucial roles in the development of depression through their activation of the mammalian target of rapamycin (mTor) pathway[21]. Our results in this study show that that at baseline higher levels of two intermediates of the BCAA biosynthesis pathway, 3-methyl-2-oxopentanoate and 4-methyl-2-oxopentanoate were correlated with more severe depressive and anxiety symptoms and a decrease in these two metabolites over the course of 12 weeks of CBT or duloxetine treatment was associated with improved symptoms. In contrast, an increase in valine with exposure to CBT or duloxetine were correlated with improvements in depressive symptoms. The decrease in these intermediates potentially may be due to increased synthesis of the downstream BCAA in response to duloxetine exposure. None of these changes were observed in the escitalopram arm suggesting that even though the drugs were more similar in their associations with metabolite alterations compared to CBT, they differed in their impact on the BCAA biosynthesis pathway.

There are several limitations to our study. The first limitation is that more insights could be gained by a dose response study. A single dose of the medications used in the experiment may not be sufficient to fully understand the effects or potential benefits of the treatment being studied. The second limitation is that we did not have catecholamines profiled. Catecholamines are a group of hormones that include metabolites like adrenaline and noradrenaline. Profiling catecholamines could provide important information about their involvement in the biochemical processes being studied; this is especially relevant because our comparisons included a SSRI and a SNRI, the latter largely associated with the catecholamine metabolic pathway. Without these data, it is challenging to fully understand the interactions and effects of the treatment. The final limitation is the inability to see the link between BCAA (branched-chain amino acids) biochemistry and other metabolites. BCAAs compromise a group of essential amino acids that play important roles in various biochemical processes in the body. However, their interactions with other metabolites, such as other amino acids or metabolic intermediates, may provide further insights into their functions and potential effects. Without studying these links, our understanding of the overall biochemical processes was limited.

## AUTHOR CONTRIBUTIONS

S.B. and S.M. did analyses of data and helped write the manuscript; B.K. and his team generated biochemical data and wrote its methods and helped with interpretation of findings; B.W.D., W.E.C. and A.J.R. helped with interpretation of findings and clinical relevance; H.S. helped with background literature searches. R.K.D. is PI for project and helped with concept development, study design, data interpretation and connecting biochemical and clinical data, and with the writing of the manuscript.

## Data Availability

All data produced in the present study are available upon reasonable request to the authors.

## ABBREVIATIONS

HRSA_14_: 14-item Hamilton Anxiety Rating Scale
HRSD_17_: 17-item Hamilton Depression Rating Scale
MDD: Major Depressive Disorder
PReDICT: Predictors of Remission in Depression to Individual and Combined Treatments study

## ACKNOWLEDGEMENTS

We acknowledge the assistance of Ms. Lisa Howerton (Duke).

## CONTRIBUTION TO THE FIELD STATEMENT

This study contributes to the field of major depressive disorder by identifying and mapping biochemical signature associated with 3 different therapies.

---

**Supplementary Figure 1:** Forest Plots Showing Baseline Association (and 95% confidence interval) of Clinical Outcomes and Metabolite Levels within each Strata.

**Supplementary Figure 2:** Forest Plots Showing 12 Weeks change and 95% confidence Interval of metabolites in response to treatment within each Strata

