## Supplementary figures and images for "Metabolomics Signatures of serotonin reuptake inhibitor (Escitalopram), serotonin norepinephrine reuptake inhibitor (Duloxetine) and Cognitive Behavior Therapy on Key Neurotransmitter Pathways in Major Depressive Disorder"

### Sup Fig-2

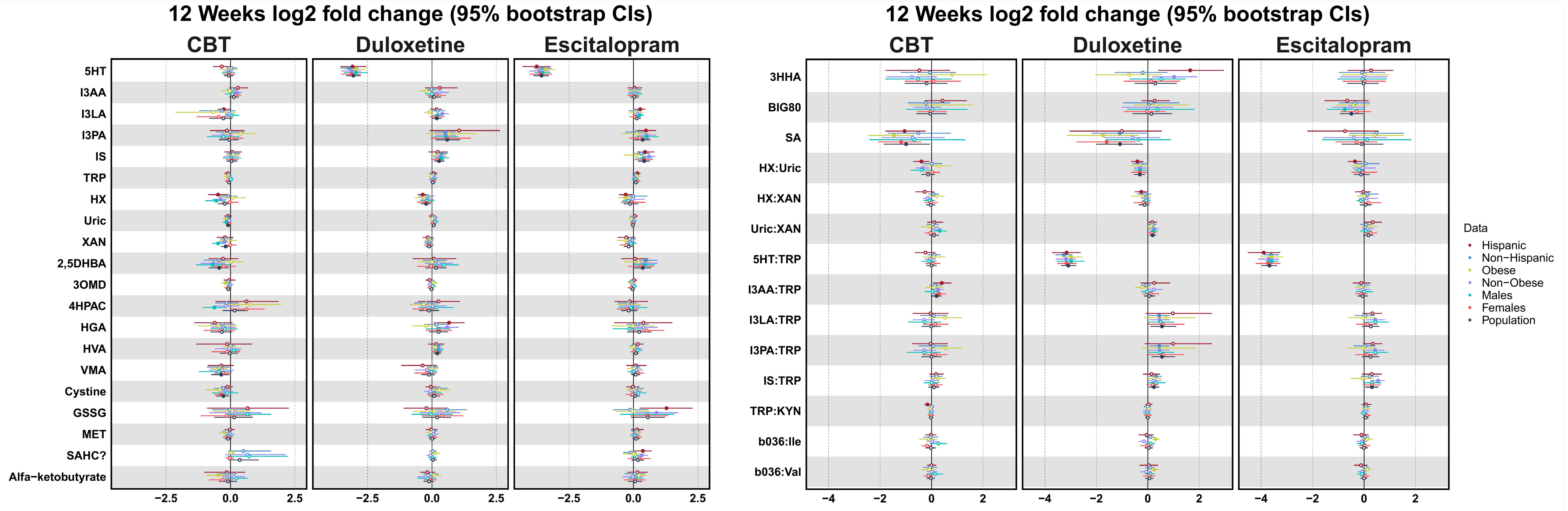

### Sup Fig- 1

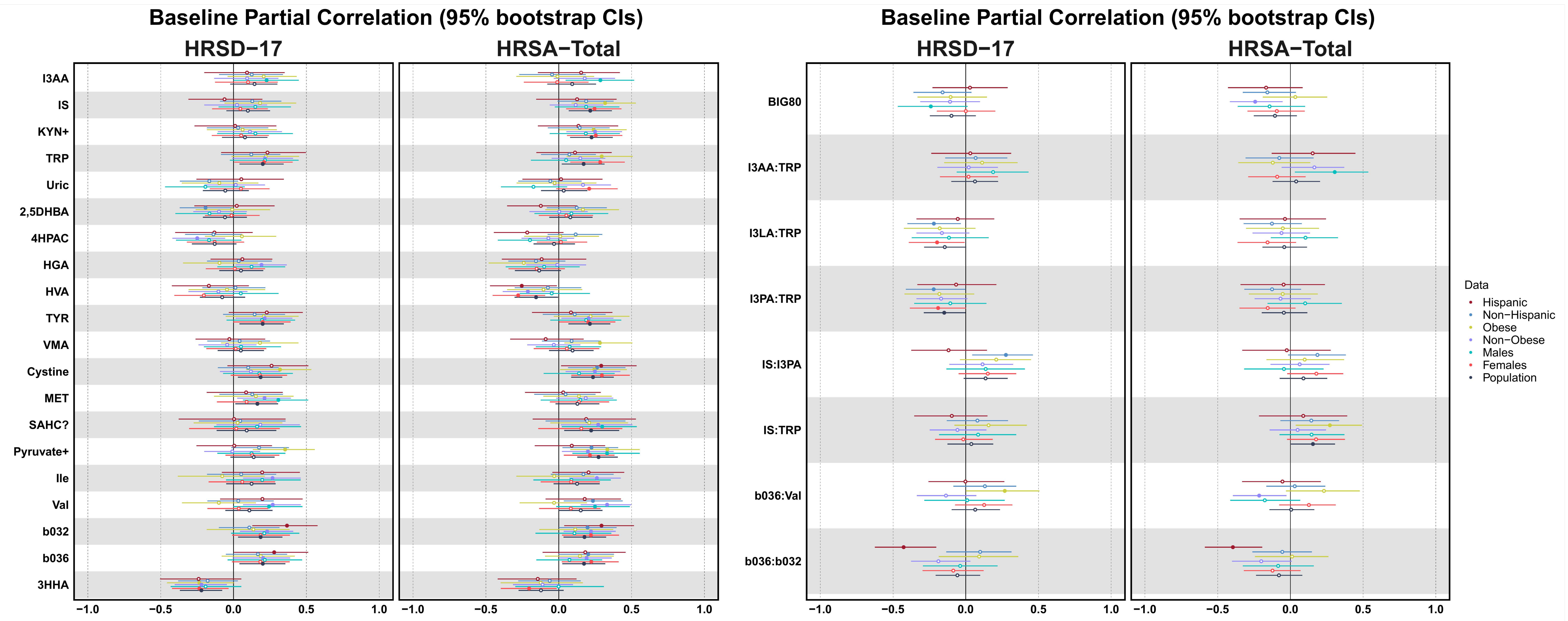
